# Physical Activity Shapes Brain Structure, Function, and the Computational Mechanisms of Cognitive Control

**DOI:** 10.64898/2025.12.12.25342075

**Authors:** Patricio Carvajal-Paredes, Joselina Davyt-Colo, Alejandra Figueroa-Vargas, María Paz Martínez-Molina, Carla Manterola, Ximena Stecher, Francisco Zamorano, Patricia Soto-Icaza, Pablo Billeke

## Abstract

**Background:** Sedentarism is prevalent and associated with poorer mental and physical health. Whether everyday physical activity (PA) maps onto computational decision strategies and brain structure/function in non-elderly adults remains unclear.

**Methods:** Seventy-one healthy adults (39 women; 18–45 years) completed the Multi-Source Interference Task (MSIT) during fMRI. PA was quantified with the short IPAQ and participants were classified as Active or Sedentary. Behavior (RT, accuracy) was analyzed with frequentist models and a hierarchical Bayesian drift–diffusion model (DDM) estimating drift rate (*v*), boundary separation (*α*), and non-decision time (τ). First-level fMRI modeled congruent/incongruent trials; group-level analyses used FLAME1 with cluster-wise FWE correction (z>3.1, p <.05). Structural MRI was processed with FreeSurfer 7.4.1 (cortical thickness, hippocampal subfields); surface-based GLMs tested group and DDM effects.

**Results:** Active participants responded faster overall; incongruent accuracy showed a speed– accuracy trade-off (accuracy increased with slower RTs), with a significant RT × group interaction. In the DDM, boundary separation (*α*) was higher in sedentary individuals (greater caution), whereas drift rate (*v*) differed by sex (males > females). Structurally, the active group showed larger left hippocampal subfields and thicker cortex in posterior temporal & anterior cingulate regions that are negatively related to *α*. Functionally, boundary-related BOLD modulation encompassed fusiform, posterior cingulate, superior temporal cortex, and SMA; Active > Sedentary contrasts highlighted left occipital and inferior parietal lobe.

**Conclusions:** Everyday PA aligns with lower decision thresholds, select structural advantages, and more efficient task-engaged networks, suggesting PA fosters adaptive, resource-efficient cognitive control. These mechanistic links support PA-based strategies to mitigate risks of sedentarism.

## 1. Introduction

Modern lifestyle patterns have increasingly shifted toward behaviors that seriously threaten public health. Sedentary behavior has emerged as a major concern, given its strong association with cardiovascular disease, type 2 diabetes, obesity, and certain cancers^1,2^. In Latin America, sedentarism rates are alarmingly high. In Chile, national surveys indicate that about 86.7% of adults reported no leisure-time physical activity in the past month^3^. This widespread sedentarism contributes not only to a growing obesity epidemic^4^ but also to increasing public health costs and diminished overall well-being^5,6^.

Despite extensive evidence on the benefits of an active lifestyle, global levels of physical inactivity continue to rise, and most interventions achieve only modest, short-lived changes in behavior^7, 8^. This suggests that powerful habitual and contextual forces sustain sedentary lifestyles and that single-component interventions are unlikely to be sufficient. In this context, understanding sedentarism from multiple perspectives is crucial for designing more effective, multidimensional strategies.^9^ One often-neglected dimension is the interplay between physical activity and cognition. Regular physical exercise promotes improvements in executive functions such as attention, memory, and self-regulation ^10,11^, which may in turn support the consolidation of physical activity as a stable habit^10,11^. Recent frameworks propose that this bidirectional relationship is mediated by neurocognitive systems—including reward, executive control, interoception, and stress-regulation networks-that can form either *virtuous or vicious cycle*^11,10,11^. Yet, the specific neurocognitive computations and large-scale control networks that support or hinder the transition from sedentary to active lifestyles remain poorly understood.

In addition, sedentary lifestyles have been consistently linked to adverse mental health outcomes, including higher rates of depression and anxiety^12^, whereas regular physical activity is associated with improvements in mood regulation, sleep quality, and cognitive performance^13^. Neuroimaging research further shows that physical activity relates to structural and functional brain adaptations, such as increased hippocampal volume and greater gray matter density in prefrontal and limbic regions^11,14^. These regions are central nodes in the cognitive control network—a set of executive functions encompassing attention regulation, inhibitory control, working memory, and cognitive flexibility ^15–26^. Cognitive control relies on the coordinated activity of frontal and parietal cortices^19,23,27,28^ and on integrative hubs such as the anterior cingulate cortex and insula, which contribute to conflict detection, decision-making, and emotion regulation^29–32^. The integrity and functional connectivity of these regions correlate with physical activity levels^33^, suggesting that sedentarism may compromise neural efficiency in high-demand cognitive tasks^34^. However, while the neural architecture of cognitive control has been well characterized^35^, it remains unclear how physical activity shapes the specific computations implemented within these networks.

Computational frameworks—such as drift diffusion and Bayesian decision models—capture latent components of cognition, including evidence accumulation, decision thresholds, and adaptive control dynamics, that cannot be inferred from neural activation alone^20,26,36^. This perspective is crucial to understand how physical activity influences decision-making strategies and cognitive efficiency beyond gross measures of activation or structure. Moreover, while several studies have examined physical activity’s effects on cognitive aging^37–39^, fewer have addressed its role in shaping decision-making and brain structure–function relationships in younger adults. By integrating neuroimaging and computational modeling, the present study aims to elucidate the neurocognitive impact of sedentarism.

Engaging in sustained physical activity requires the continuous coordination of perceptual, motor, and executive systems under variable sensory and environmental demands, effectively functioning as a natural and dynamic form of cognitive training. Consequently, physically active individuals are expected to display neurobiological and cognitive profiles characterized by greater efficiency and adaptability than their sedentary counterparts. Within this framework, we hypothesize that regular physical activity induces neuroplastic adaptations that optimize the brain’s structural integrity, functional efficiency, and computational dynamics. Specifically, by examining neurobiological and neurocomputational differences between physically active and sedentary adults during a cognitive control task, we predict that active individuals will exhibit more efficient cognitive processing, reflected in distinct drift diffusion model (DDM) profiles and corresponding structural and functional brain patterns.

## 2. Materials and Methods

### 2.1 Participants

A total of 71 healthy volunteers (39 women), aged 18–45 years (mean age = 27.5, SD = 6.22), participated in functional Magnetic Resonance Imaging (fMRI) scanning. All participants had normal or corrected-to-normal vision, no history of neurological injury, psychiatric diagnoses, or use of psychotropic medications. Informed consent was obtained from all participants. The study protocol was approved by the Scientific Ethics Committee of the Faculty of Medicine, Clínica Alemana de Santiago – Universidad del Desarrollo (Approval ID: 2023-119).

### 2.2 Physical Activity Assessment

Participants were classified as physically active or sedentary using the short form of the International Physical Activity Questionnaire (IPAQ; Craig et al., 2003). Activity levels were computed in MET-minutes/week (Table 1).

**Table 1.** Distribution by physical activity condition and Sex.

| Group | Count | Mean | Std Dev | Min | 25% | 50% | 75% | Max |
| --- | --- | --- | --- | --- | --- | --- | --- | --- |
| Active (A) | 43 | 3530 | 2513 | 639 | 1677 | 2628 | 4632 | 11093 |
| Sedentary (S) | 28 | 392 | 144 | 99 | 277 | 396 | 519 | 582 |
| Female (F) | 39 | 1699 | 1796.2 | 169.5 | 417 | 996 | 2514 | 8076 |
| Male (M) | 32 | 3015 | 3004.4 | 99 | 547.5 | 2305.5 | 4596 | 11093 |

### 2.3 Experimental Task

Participants performed the (Multi-Source Interference Task, MSIT^40^), during fMRI acquisition (Fig. 1A). In this task, subjects were shown triplets of digits (e.g., “1 0 0”) and instructed to identify the unique digit by pressing the corresponding key. Trials were either congruent (e.g., position and identity match) or incongruent (e.g., “2 3 3”, where position and identity mismatch), with 50% of trials introducing conflict. Each trial lasted 1000 ms., with inter-stimulus intervals varying between 1500 and 3000 ms. This task assesses core executive functions like Inhibitory control, Selective attention, Cognitive flexibility, and Conflict resolution^19,27,40^

**Figure 1.**
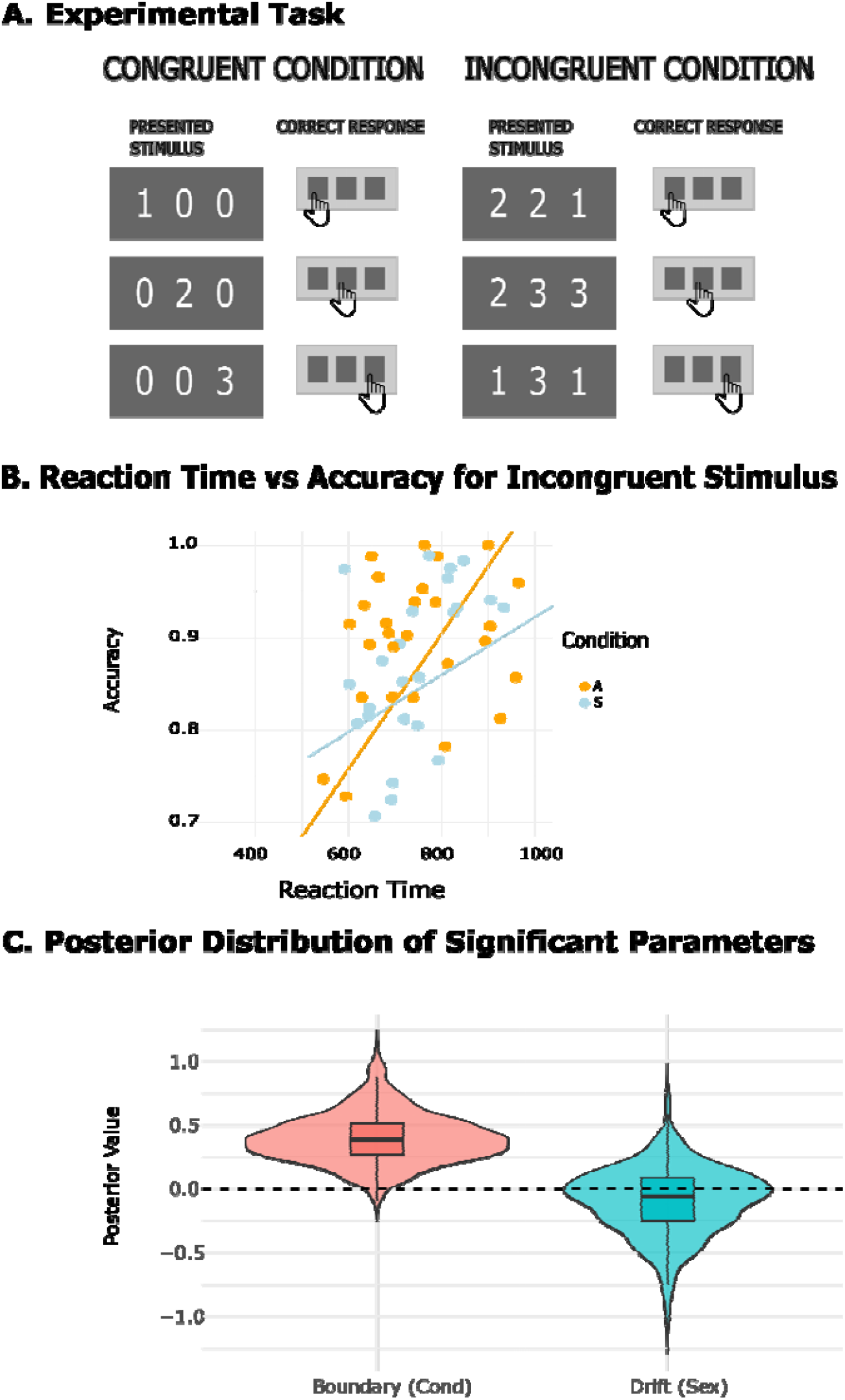
MSIT Task. Reaction Time vs Accuracy for Incongruent Stimulus & violin graph of parameters distributions. **(A)** MSIT schematic with examples of congruent and incongruent trials and the correct key for each stimulus. **(B)** Relationship between Reaction Time (ms) and Accuracy in incongruent trials (dots = participants; lines = linear fits by group (A = active, S = sedentary). **(C)** Posterior distributions (violin + box) for parameters with 95% credible intervals excluding 0. Shown: Drift (Sex) and Boundary (Condition); dashed line marks 0 on the posterior scale.

### 2.4 Behavioral Data Analysis

Behavioral data (reaction times and accuracy) were first analyzed using frequentist statistics to examine interactions between *condition* (active vs. sedentary), *stimulus type* (congruent vs. incongruent), and *sex*. Parametric and non-parametric tests were used as appropriate to compare group-level distributions and evaluate significance.

To further explore possible trade-offs between response speed and accuracy, we examined the relationship between reaction time and accuracy in the incongruent condition (Figure 1b). These behavioral metrics allow us to investigate whether participants modulate their speed and precision in response to cognitive demands (equation 4) —a phenomenon commonly referred to as the speed–accuracy trade-off, and has been a key processing modulating by cognitive control in conflict situations ^19,23^

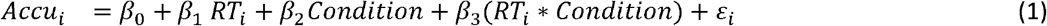

### 2.5 Bayesian Modeling with Drift Diffusion Model (DDM)

To model decision-making strategies, we applied a hierarchical Bayesian drift-diffusion model (DDM). The DDM decomposes decisions into several latent components: the non-decision time (*Ta*), reflecting sensory and motor latencies, and the drift rate (*v*), indexing the efficiency of evidence accumulation. ^41,42^ The DDM was implemented in JAGS v4.3.0 via RStudio v4.1.0, using Markov Chain Monte Carlo (MCMC) sampling (3 chains with distinct initial values). Posterior distributions were assessed for convergence and precision. Stimulus type (congruent/incongruent), sex, and condition (active/sedentary) were modeled as fixed effects and interactions.

The core model equation:

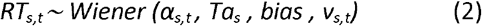

Where RT_t,s_ represent observed response time for trial t and participant s, *α* the boundary separation, *v* the drift rate, *Ta* the non-decision time, *stim* the stimulus congruency (0 or 1) and bias *the* starting bias (set as *0*.*5)*.

Linear mixed-effects modeling accounted for:

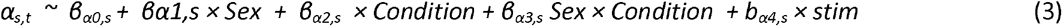

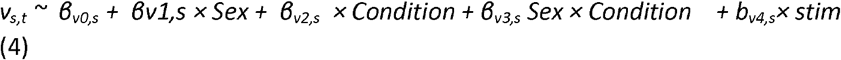

Posterior distributions of each parameter were extracted for each subject and used as regressors in subsequent neuroimaging analyses, enabling brain-behavior mapping of cognitive control strategies.

### 2.6 Neuroimaging Acquisition

MRI data were acquired at Clínica Alemana de Santiago using a 3 Tesla Siemens Skyra scanner equipped with a 20-channel head and neck coil. For each participant, both high-resolution anatomical and functional images were collected. Structural images were acquired using a T1-weighted sequence with an isotropic voxel size of 1 × 1 × 1 mm. This sequence included 176 slices, a field of view (FoV) of 256 mm, a repetition time (TR) of 2.53 seconds, and an echo time (TE) of 2.2 milliseconds. These parameters allowed for precise anatomical mapping of the brain, essential for subsequent registration and normalization of functional data.

Functional imaging during the MSIT task was performed using a T2*-weighted echo-planar imaging (EPI) sequence. Acquisition parameters were as follows: voxel size of 2.97 × 2.97 × 4 mm, 38 slices per volume, a FoV of 220 mm, TR of 2.21 seconds, and TE of 30 milliseconds. This setup enabled the capture of blood-oxygen-level-dependent (BOLD) signal changes associated with cognitive task performance. Visual stimuli were delivered using Presentation® software (Neurobehavioral Systems) through the MRI-compatible VisuaStimDigital system (Resonance Technology Inc.), and participants provided responses via button presses using an integrated response device while remaining inside the scanner.

### 2.7 fMRI Preprocessing and First-Level Analysis

Functional MRI data were preprocessed using the FMRIB Software Library (FSL, version 6.0.7.6). The preprocessing pipeline included motion correction using MCFLIRT, slice-timing correction, brain extraction via BET, and spatial smoothing with a Gaussian kernel of 6 mm full-width at half maximum (FWHM). A high-pass temporal filter with a sigma of 60.0 seconds was applied to remove low-frequency drifts, followed by prewhitening to correct for temporal autocorrelation.

The preprocessed time series were analyzed using a general linear model (GLM), in which separate explanatory variables (EVs) were defined for congruent and incongruent trials. Each EV was convolved with a canonical double-gamma hemodynamic response function. Additionally, motion parameters were included as nuisance regressors to account for residual movement artifacts.

To allow for group-level analysis, individual contrast maps were normalized to the standard MNI152 template in a two-step registration process. First, each participant’s functional images were aligned to their respective T1-weighted structural images using linear registration with six degrees of freedom. Then, the anatomical scans were registered to MNI152 space using affine transformation followed by non-linear registration using FNIRT. Second-level group analyses were performed using FLAME1 (FMRIB’s Local Analysis of Mixed Effects), which accounts for both within- and between-subject variance. Statistical thresholding was applied using a cluster-forming threshold of z > 3.1, with family-wise error (FWE) correction at p < 0.05, in line with current recommendations for robust and reliable cluster inference in fMRI research^43^.

### 2.8 Structural MRI Analysis

Structural MRI data were analyzed using FreeSurfer version 7.4.1, a software suite for the automated segmentation of brain structures and estimation of cortical thickness^44^. The preprocessing pipeline included intensity normalization, skull stripping, and the segmentation of white and gray matter tissues. Cortical thickness was computed as the distance between the pial surface and the white matter boundary at each vertex across the cortical mantle. Volumetric segmentation of subcortical regions was conducted using the asegstats2table function, which allowed for the extraction of region-of-interest (ROI) volumes for statistical analysis.

Further structural characterization involved detailed segmentation of thalamic nuclei and hippocampal subfields. Thalamic nuclei were segmented using the segment *ThalamicNuclei*.*sh* tool, which relies on a probabilistic atlas and high-resolution T1-weighted volumes. Hippocampal and amygdalar subfields were segmented using the -hippo-subfields flag in the recon-all pipeline. This approach utilizes ultra-high resolution ex vivo data to delineate fine-grained structures such as CA1, CA2/3, CA4, the dentate gyrus (DG), subiculum, and individual amygdalar nuclei.

Surface-based group analyses of cortical thickness were conducted using the SurfStat toolbox implemented in MATLAB. For this analysis, a matrix of cortical thickness values (vertices × subjects) was constructed. A GLM was then fitted, including predictors for group (active vs. sedentary), sex, and their interaction. Statistical contrasts were defined to assess differences in cortical thickness between groups. To correct for multiple comparisons across the cortical surface, Random Field Theory (RFT) was applied. Finally, statistically significant results were visualized by projecting the findings onto inflated cortical surfaces, allowing for intuitive and anatomically informed interpretation.

## 3. Results

### 3.1 Behavioral Performance

All subjects demonstrated adequate performance during the MSIT task (Figure 1A). Accuracy (ACCU) in both groups was above chance level (33%), with a mean of 0.80 (SD = 0.20, range = 0.44–1.00) in the Active (A) group and 0.92 (SD = 0.06, range = 0.73–0.99) in the Sedentary (S) group. There was no significant difference in accuracy between groups (Mann-Whitney U = 464, p = 0.11). Reaction times (RT) were slower in the Sedentary group (mean = 631.67 ms, SD = 131.74, range = 441.98–986.16 ms) compared to the Active group (mean = 554.87 ms, SD = 122.18, range = 353.92–820.70 ms). This difference was statistically significant (Mann-Whitney U = 23,502,490, p < 0.001), suggesting increased cognitive demand or conflict in the Sedentary group.

Reaction times were significantly slower in incongruent trials (mean = 688 ms, SD = 240) compared to congruent trials (mean = 479 ms, SD = 146), and accuracy was significantly lower in incongruent (mean = 0.73, SD = 0.44) than in congruent trials (mean = 0.98, SD = 0.13; all p < 0.001). This conflict effect was observed in both groups (p < 0.001): in the Active group, reaction times increased from 465 ms (SD = 133) to 642 ms (SD = 233) and accuracy decreased from 0.98 (SD = 0.13) to 0.62 (SD = 0.49); in the Sedentary group, reaction times increased from 495 ms (SD = 158) to 737 ms (SD = 239), and accuracy decreased from 0.99 (SD = 0.12) to 0.85 (SD = 0.36).

While physically active participants responded faster on average, sedentary participants exhibited greater overall accuracy. Considering the stimulus type, accuracy remained high in congruent trials for both groups, while greater divergence in reaction times appeared during incongruent (conflict) trials. In the incongruent condition, reaction times remained slower in the Sedentary group (mean = 755.23 ms, SD = 162.92, range = 512.30–986.16 ms) than in the Active group (mean = 644.92 ms, SD = 168.99, range = 356.97–820.70 ms), with this difference reaching statistical significance (Mann-Whitney U = 391, p = 0.013). In contrast, accuracy did not show a significant difference in the incongruent trials between the Sedentary group (mean = 0.85, SD = 0.12, range = 0.73–0.99) compared to the Active group (mean = 0.62, SD = 0.38, range = 0.44–1.00) (Mann-Whitney U = 467.5, p = 0.11).

Simple linear regression models explored the interaction between accuracy and RT across conditions and stimulus type (Table 2). In incongruent trials, accuracy was significantly predicted by RT (β = 0.00073, p <.001), indicating that higher accuracy was associated with slower responses. The interaction term for condition and RT was also significant (β = −0.00042, p =.045), suggesting that the relationship between accuracy and RT differs by activity level. However, models for congruent trials yielded no significant effects or interactions (all |t| ≤ 1.55, all, ps ≥.13), implying that group differences manifest primarily in more demanding cognitive conditions.

**Table 2.**
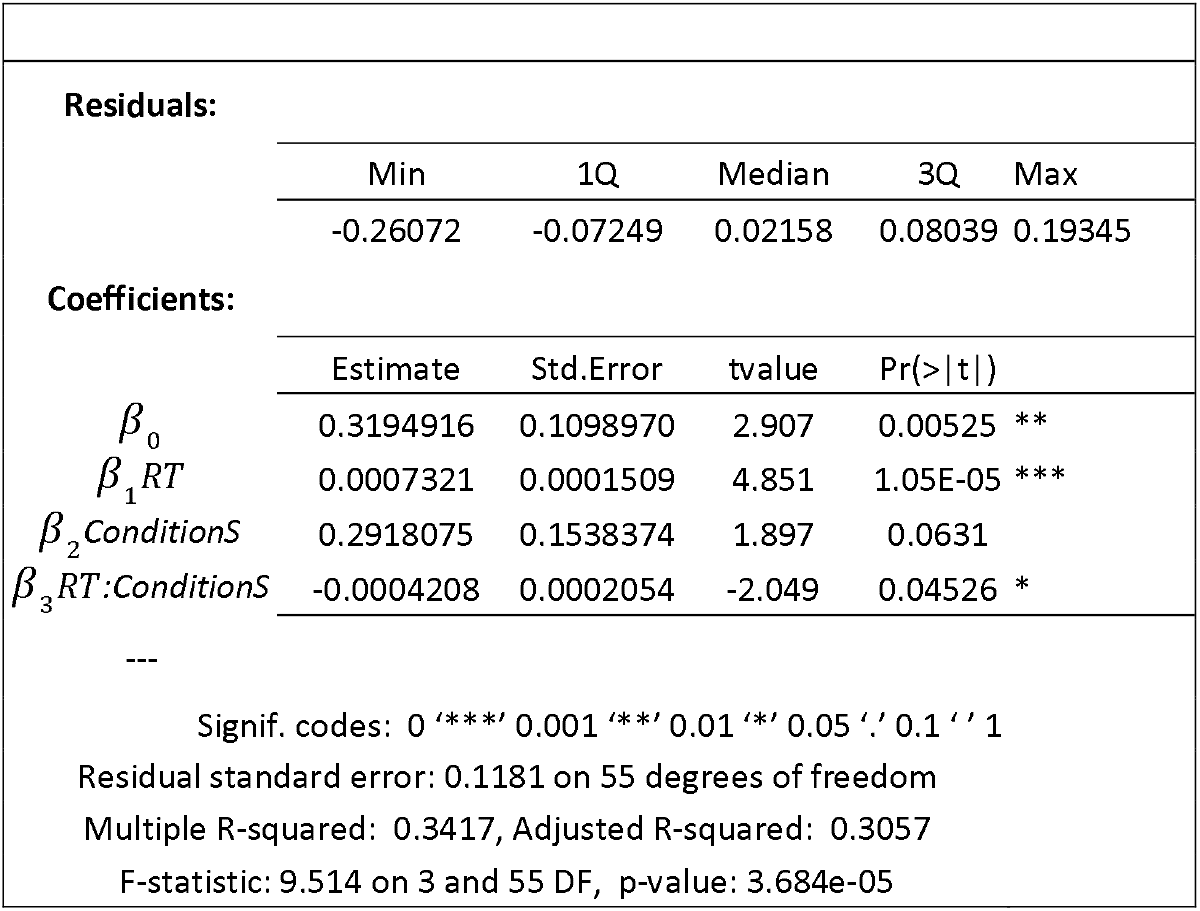
Linear Model of RT and Accu in incongruent trials.

### 3.2 Bayesian Drift Diffusion Modeling (DDM)

To gain a more mechanistic understanding of the speed–accuracy trade-off, we evaluated cognitive computations using a DDM approach^19,20,28^. Specifically, we applied a hierarchical Bayesian DDM to decompose cognitive control processes into latent computational components. The model estimated three parameters: drift rate (v), reflecting the speed of information accumulation; boundary separation (a), reflecting the speed–accuracy trade-off; and non-decision time (τ), reflecting sensory–motor processing. This type of model has been previously used in similar tasks, demonstrating good fit^19^. All parameters were composed of a baseline estimate (β0) and the effect of the conditions of interest, namely sex, sedentary/active group, and task condition (see Methods).

All parameters showed good convergence (PSRF < 1.1). Notably, the drift rate differed significantly by sex, with males accumulating evidence more rapidly, while boundary separation differed by physical activity, with sedentary individuals requiring more evidence to commit to a decision (see Table 3 and posterior parameters distributions on Figure 1c).

**Table 3.**
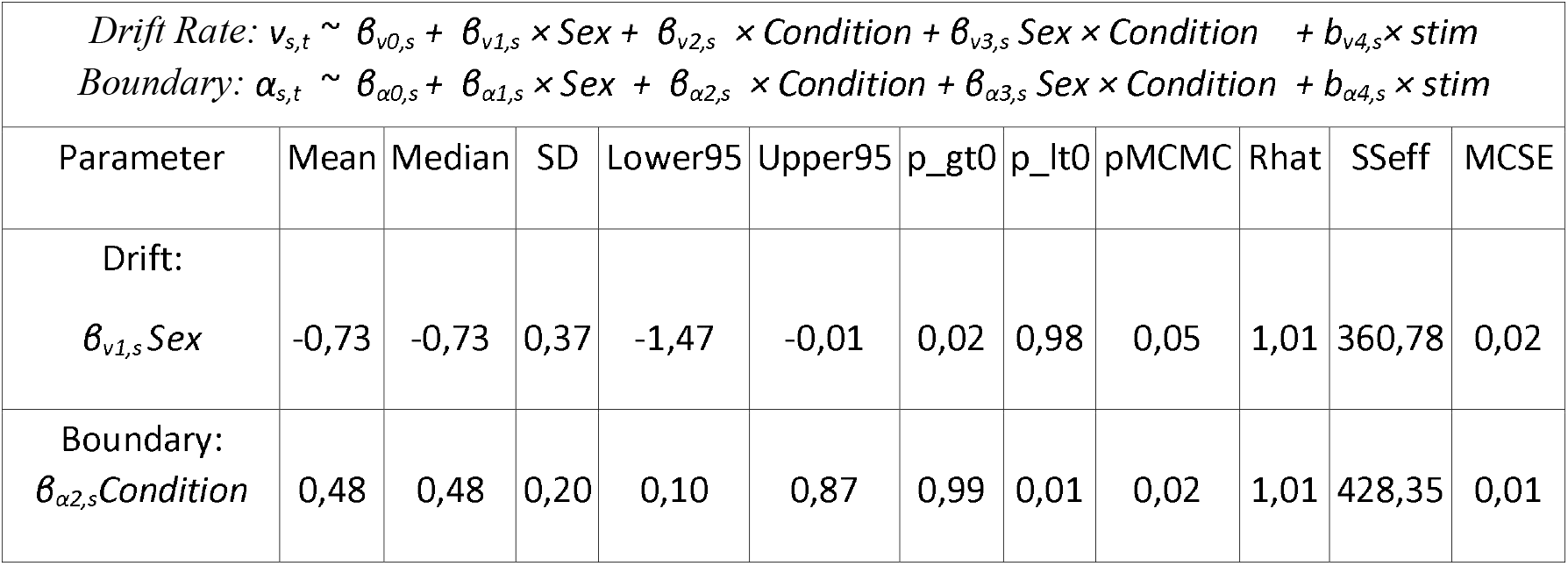
Significant DDM parameter estimates.

These behavioral parameters, estimated by the Bayesian drift diffusion mode, will be used as subject-level regressors in our neuroimaging analysesl—particularly the boundary separation, which differentiates sedentary and physically active participants—. This will allow us to test whether individual differences in decision making process are systematically associated with variations in brain structure and functional activation patterns, thereby linking computational indices of cognitive control to their neural substrates.

### 3.3 Brain Results

#### 3.3.1 Structural Brain Results. Hippocampus

We first focused on the hippocampus, given its well-established sensitivity to social and environmental influences — including stress, cognitive stimulation, and physical exercise — that can modulate its morphology and function^45,46^. To assess volumetric differences, we conducted ANOVAs controlling for sex, which revealed that physically active individuals exhibited significantly greater normalized hippocampal volumes across several subfields of the left hemisphere (see Supplementary Table 1). Consistent with these volumetric findings, shape analyses using vertex-wise morphometry showed medium-to-large effect sizes in left hippocampal regions, particularly within the CA3-head, CA4-head, and dentate gyrus (DG-body), again favoring the active group (see Figure 2a & Supplementary Table 2). These results suggest that habitual physical activity may be associated with structural preservation or enhancement in hippocampal subregions critically involved in memory and stress regulation.

**Figure 2.**
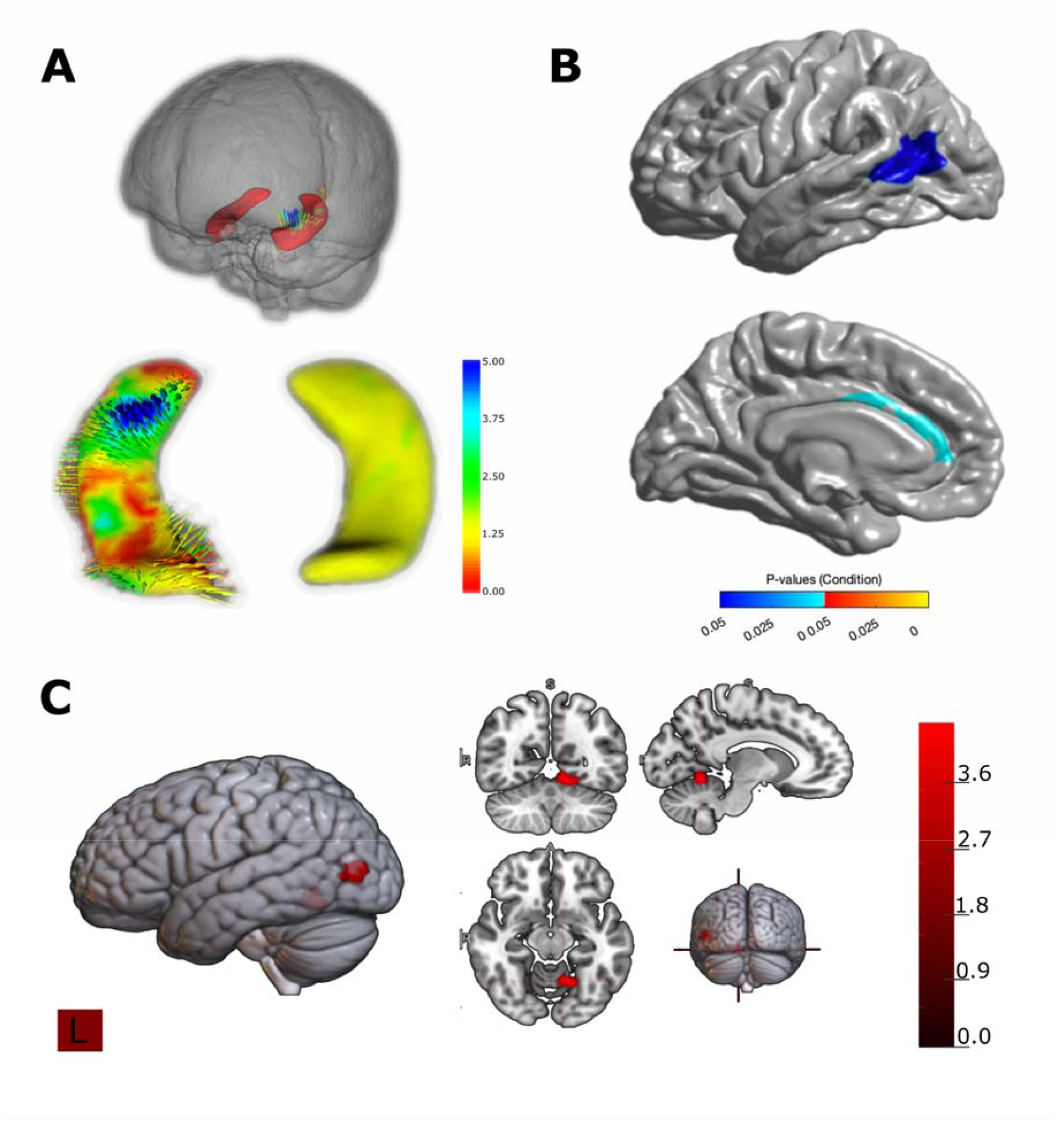
Hippocampal shape Analysis. Cortical Thickness and Functional Results. **(A)** Vertex-wise shape analysis of the hippocampus showing regions of displacement between groups. Warm colors indicate outward deformation and greater local expansion, whereas cool colors indicate inward displacement; arrows depict the direction and magnitude of the surface change. The inset shows the bilateral hippocampal location in the whole brain. **(B)** Cortical surface analysis revealing a significant cluster on the left superior temporal/supramarginal region (upper) and an extended effect along the superior temporal sulcus/middle temporal gyrus (lower). **(C)** Cluster-level analysis on the cortical surface (left) and in orthogonal views (right) confirming a focal involvement of the left superior temporal region; color bar indicates statistical values (higher values in red).

#### 3.3.2 Cortical Thickness

Building on the behavioral findings that revealed group differences in the speed–accuracy trade-off and sex-related variations in evidence accumulation, we incorporated individual estimates of the key DDM parameters — boundary separation (a) and drift rate (v), or their corresponding β coefficients — as regressors in both structural and functional analyses. This approach allowed us to identify the neurobiological substrates associated with interindividual variability in decision-making dynamics and to elucidate how cortical architecture and activity patterns relate to these computational mechanisms (e.g., ^46,47^).

A generalized linear model (GLM) was applied at each cortical vertex, including the boundary separation parameter (*a*), sex, and condition as predictors. Significant negative associations were observed between boundary separation and cortical thickness in two regions: the posterior temporal cortex (spanning the intersection of Brodmann areas 19, 22, 37, and 39) and the anterior cingulate cortex (see Figure 2b). These results indicate that individuals exhibiting higher decision thresholds—particularly those in the sedentary group—show reduced cortical thickness in regions critically involved in cognitive control and performance monitoring.

#### 3.3.3 Functional MRI Results

We analyzed task-related BOLD responses focusing on the MSIT effect, defined as the contrast between incongruent and congruent correct trials. Across all 71 participants, this contrast elicited robust activation within the canonical cognitive control network (see Supplementary table 3), including the dorsomedial prefrontal cortex, inferior frontal gyrus, anterior cingulate cortex, caudate nucleus, and fusiform and lingual gyri — consistent with prior evidence of conflict-related engagement of fronto-striatal and occipito-temporal regions.

We then examined how individual differences in the boundary separation parameter (*a*) modulated task-related BOLD activity by including this regressor in the second-level analysis (see Methods). This analysis revealed significant correlations between the boundary parameter and BOLD responses in the left fusiform gyrus, left posterior cingulate cortex, right superior temporal cortex, and supplementary motor area (see Supplementary table 4). These findings suggest that decision thresholds are neurally represented in a distributed network encompassing regions implicated in perceptual processing, attention allocation, and motor preparation.

Finally, contrasting boundary-related activity between groups revealed stronger boundary–BOLD coupling in the left occipital cortex and left inferior parietal lobule for the active group (Figure 2c). This pattern reinforces the interpretation that physically active individuals engage more efficient sensory and attentional systems when facing increased cognitive demands.

## 4. Discussion

This study provides evidence of structural and functional differences in the brains of physically active individuals compared with their sedentary counterparts. These functional and structural alterations were accompanied by differences in cognitive computational processes, indicating that such brain changes are not merely anatomical correlates but may translate into meaningful advantages in everyday behavior. Specifically, volumetric increases in left hippocampal subfields suggest a neuroprotective role of physical activity, aligning with prior research on exercise-induced neurogenesis and the maintenance of memory-critical structures.^48^ Interestingly, other lifestyle factors—such as dietary intake of long-chain polyunsaturated fatty acids—have also been linked to the integrity of hippocampal circuits, which in turn support real-life cognitive performance.^49^

These converging lines of evidence suggest that multiple modifiable behaviors may act synergistically to maintain hippocampal integrity and support cognitive resilience across the lifespan. However, unidimensional or non-personalized interventions tend to yield only modest effects on brain health and cognition, highlighting the need for more individualized, multimodal approaches.^50^ Characterizing the specific neurocognitive computations underlying these effects is a key step toward that goal. The cortical areas identified in our study include temporal regions centrally involved in speech perception, social cognitive processing, and audiovisual integration, highlighting their relevance for multimodal information processing^20,25,26,46,51^. Recent neuroimaging evidence demonstrates that cardiorespiratory fitness is positively correlated with cortical thickness in medial temporal regions associated with spatial cognition in young adults ^52^ and also with other cortical areas.^53,54^ Associations between aerobic fitness and hippocampal morphology remain inconsistent across studies—some reports describe volume increases associated with higher fitness levels or exercise interventions^55^, whereas others find no significant effects or only region-specific, modular patterns of change^56^

Our results suggest that physically active individuals may exhibit superior environmental responsiveness, which has direct implications for communication and cognitive processing. A key differentiating parameter in this study was the decision threshold (boundary) as derived from Bayesian drift diffusion modeling that has related cognitive control in this type of task^19,26^. Lower decision thresholds in active individuals suggest a more efficient cognitive system capable of faster decision-making. These findings align with prior literature linking physical activity to improved cognitive flexibility and executive control^57^. Functionally, the observed coactivation of sensory and motor-related brain regions suggests a more efficient integration of perceptual and executive systems, enhancing evidence accumulation and response execution.

Moreover, functional-activation patterns in physically active individuals frequently show greater involvement of sensory and multimodal processing areas—such as the fusiform gyrus, superior temporal cortex, and posterior cingulate—suggesting enhanced efficiency in sensory-driven processing. A recent meta-analysis of fMRI studies reported that aerobic-exercise interventions significantly alter task-evoked brain activation, notably in frontal, precuneus, thalamic and cingulate regions, supporting a widespread impact of exercise on functional activation beyond classical motor or control areas.^58^ In parallel, elevated activation in motor-preparation and control regions — such as the supplementary motor area and dorsomedial prefrontal cortex — highlights improved readiness and executive control in active participants. Consistent with these findings, an study found that habitual physical activity enhances prefrontal cortex engagement during implicit motor learning tasks, linking everyday fitness to more efficient motor-cognitive integration.^59^

These findings may be interpreted through the framework of cognitive reserve, which posits that sustained physical activity contributes to more resilient and efficient neural networks^60^. The increased hippocampal volumes, greater cortical thickness, and heightened activation in task-relevant regions observed in active individuals may reflect the cumulative cognitive stimulation afforded by regular physical exercise. However, additional factors such as nutrition, socioeconomic status, and genetic predispositions may also influence these outcomes. Future research should pursue longitudinal designs to explore causality and examine whether sustained physical activity induces structural and functional brain changes over time. Investigating the differential impact of aerobic versus anaerobic training could further elucidate which modalities are most beneficial for brain health.

While comprehensive, this study is not without limitations. The correlational design precludes causal inferences, and the sample size, though adequate for neuroimaging, may limit generalizability. Age-related effects were not deeply explored ^61–63^, and lifestyle factors such as diet or stress were not systematically controlled.^64^ Despite the use of state-of-the-art neuroimaging techniques, the complexity of brain organization warrants caution when interpreting activation patterns. These findings should be integrated with complementary methodological approaches to strengthen mechanistic inferences. ^25,26,28,49^

The public health implications of these findings are substantial. Promoting regular physical activity may not only prevent chronic illnesses but also enhance cognitive function and brain integrity across the lifespan ^64,65^. These results support the inclusion of movement-based interventions in schools and workplaces, particularly in systems where opportunities for physical activity are minimal. In Chile, for instance, students receive only two hours of physical education per week, and school staff often lack institutional support for integrating physical activity into their routines.

The use of advanced neuroimaging tools such as MRI, fMRI, and Bayesian modeling in this study allowed for a nuanced understanding of how physical activity modulates neural efficiency. Identifying specific brain regions affected by sedentary behavior opens pathways for targeted interventions to mitigate cognitive decline and promote brain resilience.^66^

## 5. Conclusion

This study demonstrates that regular physical activity is associated with structural and functional brain advantages, including increased cortical thickness, and enhanced hippocampal subfield volumes. These neural differences were accompanied by more efficient cognitive processing, reflected in lower decision thresholds, improved activation patterns, and optimized connectivity across sensory, attentional, and motor networks. Although causal conclusions cannot be drawn, the findings highlight the potential of physical activity to support brain health and cognitive resilience, underscoring its relevance for public health efforts aimed at mitigating the consequences of sedentary lifestyles.

## Supporting information

Supplementary_Material.docx

## Data Availability

All data produced in the present study are available upon reasonable request to the authors

## Acknowledgements

This work was supported by FONDECYT 1251073, 1211227, Clinica Alemana de Santiago

## Author Contributions

PhD Patricio Carvajal-Paredes conceptualized the study as part of a doctoral thesis, designed the methodology, recruited participants, performed the MRI investigation, coordinated study logistics and clinical administration (Project Administration), performed data analysis (Formal Analysis), and wrote the original draft. PhD Pablo Billeke provided supervision and guidance for the Bayesian formal analysis and contributed to the final writing – review & editing. PhD Patricia Soto-Icaza contributed to the conceptualization of the study and to the writing – review & editing of the manuscript. PhD Francisco Zamorano performed specific formal analysis on structural neuroimaging data. MD Ximena Stecher provided clinical reports and resources (MRI reporting). MD Carla Manterola provided expertise in clinical investigation (neuroimaging findings reporting). PhD Alejandra Figueroa-Vargas was responsible for the visualization and writing – review & editing. PhD Maria Paz Martínez-Molina contributed to the writing – review & editing manuscript. PhD(c) Joselina Davyt performed formal analysis related to the initial exploratory analysis.

## References

1. Booth FW, Roberts CK, Laye MJ. Comprehensive Physiology. Compr Physiol 2013;2:1143–211. 10.1002/cphy.c110025.

2. Tremblay MS, Aubert S, Barnes JD, Saunders TJ, Carson V, Latimer-Cheung AE, et al. Sedentary Behavior Research Network (SBRN) – Terminology Consensus Project process and outcome. Int J Behav Nutr Phys Act 2017;14:75. 10.1186/s12966-017-0525-8.

3. Chile M de S. Tercera Encuesta Nacional de Salud (ENS) 2016-2017 [Internet]. 2018 Jan. Available from: https://epi.minsal.cl/wpcontent/uploads/2017/12/2017.21.07_pdf.primeros.resultados.pdf

4. Livingston G, Huntley J, Liu KY, Costafreda SG, Selbæk G, Alladi S, et al. Dementia prevention, intervention, and care: 2024 report of the Lancet standing Commission. Lancet 2024;404:572–628. 10.1016/s0140-6736(24)01296-0.

5. Ding D, Lawson KD, Kolbe-Alexander TL, Finkelstein EA, Katzmarzyk PT, Mechelen W van, et al. The economic burden of physical inactivity: a global analysis of major non-communicable diseases. Lancet 2016;388:1311–24. 10.1016/s0140-6736(16)30383-x.

6. Pratt M, Norris J, Lobelo F, Roux L, Wang G. The cost of physical inactivity: moving into the 21st century. Br J Sports Med 2014;48:171. 10.1136/bjsports-2012-091810.

7. Guthold R, Stevens GA, Riley LM, Bull FC. Worldwide trends in insufficient physical activity from 2001 to 2016: a pooled analysis of 358 population-based surveys with 1·9 million participants. Lancet Glob Heal 2018;6:e1077–86. 10.1016/s2214-109x(18)30357-7.

8. Simpson A, Beauchamp MR, Dimmock J, Willis C, Jackson B. Health behaviour change: Theories, progress, and recommendations for the next generation of physical activity research. Psychol sport Exerc 2025;80:102918. 10.1016/j.psychsport.2025.102918.

9. Casado-Robles C, Viciana J, Guijarro-Romero S, Mayorga-Vega D. Effects of Consumer-Wearable Activity Tracker-Based Programs on Objectively Measured Daily Physical Activity and Sedentary Behavior Among School-Aged Children: A Systematic Review and Meta-analysis. Sports Med - Open 2022;8:18. 10.1186/s40798-021-00407-6.

10. Audiffren M, André N. The exercise–cognition relationship: A virtuous circle. J Sport Heal Sci 2019;8:339–47. 10.1016/j.jshs.2019.03.001.

11. Stillman CM, Erickson KI. Physical activity as a model for health neuroscience. Ann N York Acad Sci 2018;1428:103–11. 10.1111/nyas.13669.

12. Schuch FB, Vancampfort D, Firth J, Rosenbaum S, Ward PB, Silva ES, et al. Physical Activity and Incident Depression: A Meta-Analysis of Prospective Cohort Studies. Am J Psychiatry 2018;175:631–48. 10.1176/appi.ajp.2018.17111194.

13. Piercy KL, Troiano RP, Ballard RM, Carlson SA, Fulton JE, Galuska DA, et al. The Physical Activity Guidelines for Americans. JAMA 2018;320:2020. 10.1001/jama.2018.14854.

14. Erickson KI, Voss MW, Prakash RS, Basak C, Szabo A, Chaddock L, et al. Exercise training increases size of hippocampus and improves memory. Proc Natl Acad Sci 2011;108:3017–22. 10.1073/pnas.1015950108.

15. Levy R. The prefrontal cortex: from monkey to man. Brain 2023;147:794–815. 10.1093/brain/awad389.

16. Kenwood MM, Kalin NH, Barbas H. The prefrontal cortex, pathological anxiety, and anxiety disorders. Neuropsychopharmacol 2022;47:260–75. 10.1038/s41386-021-01109-z.

17. Soltani A, Koechlin E. Computational models of adaptive behavior and prefrontal cortex. Neuropsychopharmacol 2021:1–14. 10.1038/s41386-021-01123-1.

18. Haber SN, Liu H, Seidlitz J, Bullmore E. Prefrontal connectomics: from anatomy to human imaging. Neuropsychopharmacol 2021:1–21. 10.1038/s41386-021-01156-6.

19. Martínez-Molina MP, Valdebenito-Oyarzo G, Soto-Icaza P, Zamorano F, Figueroa-Vargas A, Carvajal-Paredes P, et al. Lateral prefrontal theta oscillations causally drive a computational mechanism underlying conflict expectation and adaptation. Nat Commun 2024;15:9858. 10.1038/s41467-024-54244-8.

20. Kausel L, Figueroa-Vargas A, Zamorano F, Stecher X, Aspé-Sánchez M, Carvajal-Paredes P, et al. Patients recovering from COVID-19 who presented with anosmia during their acute episode have behavioral, functional, and structural brain alterations. Sci Rep 2024;14:19049. 10.1038/s41598-024-69772-y.

21. Kausel L, Zamorano F, Billeke P, Sutherland ME, Alliende MI, Larrain-Valenzuela J, et al. Theta and alpha oscillations may underlie improved attention and working memory in musically trained children. Brain Behav 2024;14:e3517. 10.1002/brb3.3517.

22. Figueroa-Vargas A, Cárcamo C, Henríquez-Ch R, Zamorano F, Ciampi E, Uribe-San-Martin R, et al. Frontoparietal connectivity correlates with working memory performance in multiple sclerosis. Sci Rep 2020;10:9310. 10.1038/s41598-020-66279-0.

23. Zamorano F, Kausel L, Albornoz C, Lavin C, Figueroa-Vargas A, Stecher X, et al. Lateral Prefrontal Theta Oscillations Reflect Proactive Cognitive Control Impairment in Males With Attention Deficit Hyperactivity Disorder. Front Syst Neurosci 2020;14:37. 10.3389/fnsys.2020.00037.

24. Figueroa-Vargas A, Góngora B, Alonso MF, Ortega A, Soto-Fernández P, Z-Rivera L, et al. The effect of a cognitive training therapy based on stimulation of brain oscillations in patients with mild cognitive impairment in a Chilean sample: study protocol for a phase IIb, 2 × 3 mixed factorial, double-blind randomised controlled trial. Trials 2024;25:1–14. 10.1186/s13063-024-07972-7.

25. Figueroa-Vargas A, Navarrete-Caro S, Cárcamo C, Ciampi E, Vásquez-Torres M, Soler B, et al. White matter volume and microstructural integrity are associated with fatigue in relapsing multiple sclerosis. Sci Rep 2025;15:16417. 10.1038/s41598-025-01465-6.

26. Figueroa-Vargas A, Kausel L, Stecher X, Zamorano F, Aspé-Sánchez M, Carvajal-Paredes P, et al. Coupling Between Neural Oscillations and White Matter Integrity Reveals Cognitive Computational Profiles Following COVID-19. 2025. 10.21203/rs.3.rs-7538742/v1.

27. Zamorano F, Billeke P, Kausel L, Larrain J, Stecher X, Hurtado JM, et al. Lateral prefrontal activity as a compensatory strategy for deficits of cortical processing in Attention Deficit Hyperactivity Disorder. Sci Rep-uk 2017;7:7181. 10.1038/s41598-017-07681-z.

28. Valdebenito-Oyarzo G, Martínez-Molina MP, Soto-Icaza P, Zamorano F, Figueroa-Vargas A, Larraín-Valenzuela J, et al. The parietal cortex has a causal role in ambiguity computations in humans. PLOS Biol 2024;22:e3002452. 10.1371/journal.pbio.3002452.

29. Botvinick MM. Conflict monitoring and decision making: Reconciling two perspectives on anterior cingulate function. Cogn, Affect, Behav Neurosci 2007;7:356–66. 10.3758/cabn.7.4.356.

30. Lamm C, Singer T. The role of anterior insular cortex in social emotions. Brain Struct Funct 2010;214:579–91. 10.1007/s00429-010-0251-3.

31. Billeke P, Ossandon T, Perrone-Bertolotti M, Kahane P, Bastin J, Jerbi K, et al. Human Anterior Insula Encodes Performance Feedback and Relays Prediction Error to the Medial Prefrontal Cortex. Cereb Cortex 2020;30:4011–25. 10.1093/cercor/bhaa017.

32. Lavín C, Soto-Icaza P, López V, Billeke P. Another in need enhances prosociality and modulates frontal theta oscillations in young adults. Front Psychiatry 2023;14:1160209. 10.3389/fpsyt.2023.1160209.

33. Voss MW, Carr LJ, Clark R, Weng T. Revenge of the “sit” II: Does lifestyle impact neuronal and cognitive health through distinct mechanisms associated with sedentary behavior and physical activity? Ment Heal Phys Act 2014;7:9–24. 10.1016/j.mhpa.2014.01.001.

34. Zhao Y, Li Y, Wang L, Song Z, Di T, Dong X, et al. Physical Activity and Cognition in Sedentary Older Adults: A Systematic Review and Meta-Analysis. J Alzheimer’s Dis 2022;87:957–68. 10.3233/jad-220073.

35. Menon V, D’Esposito M. The role of PFC networks in cognitive control and executive function. Neuropsychopharmacol 2021;47:1–14. 10.1038/s41386-021-01152-w.

36. Ratcliff R. A diffusion model account of response time and accuracy in a brightness discrimination task: Fitting real data and failing to fit fake but plausible data. Psychon B Rev 2002;9:278–91. 10.3758/bf03196283.

37. Colcombe S, Kramer AF. Fitness Effects on the Cognitive Function of Older Adults. Psychol Sci 2002;14:125–30. 10.1111/1467-9280.t01-1-01430.

38. Falck RS, Davis JC, Liu-Ambrose T. What is the association between sedentary behaviour and cognitive function? A systematic review. Br J Sports Med 2017;51:800–11. 10.1136/bjsports-2015-095551.

39. Liu-Ambrose T, Nagamatsu LS, Graf P, Beattie BL, Ashe MC, Handy TC. Resistance Training and Executive Functions: A 12-Month Randomized Controlled Trial. Arch Intern Med 2010;170:170–8. 10.1001/archinternmed.2009.494.

40. Bush G, Shin LM. The Multi-Source Interference Task: an fMRI task that reliably activates the cingulo-frontal-parietal cognitive/attention network. Nat Protoc 2006;1:308–13. 10.1038/nprot.2006.48.

41. Wagenmakers E-J, Ratcliff R, Gomez P, McKoon G. A diffusion model account of criterion shifts in the lexical decision task. J Mem Lang 2008;58:140–59. 10.1016/j.jml.2007.04.006.

42. Ratcliff R, Thapar A, Gomez P, McKoon G. A Diffusion Model Analysis of the Effects of Aging in the Lexical-Decision Task. Psychol Aging 2004;19:278–89. 10.1037/0882-7974.19.2.278.

43. Eklund A, Nichols TE, Knutsson H. Cluster failure: Why fMRI inferences for spatial extent have inflated false-positive rates. Proc Natl Acad Sci 2016;113:7900– 5. 10.1073/pnas.1602413113.

44. Dale AM, Fischl B, Sereno MI. Cortical Surface-Based Analysis. Neuroimage 1999;9:179–94. 10.1006/nimg.1998.0395.

45. McEwen BS, Gianaros PJ. Stress- and Allostasis-Induced Brain Plasticity. Annu Rev Med 2011;62:431–45. 10.1146/annurev-med-052209-100430.

46. Billeke P, Ossandon T, Stockle M, Perrone-Bertolotti M, Kahane P, Lachaux J-P, et al. Brain state-dependent recruitment of high-frequency oscillations in the human hippocampus. Cortex 2017;94:87–99. 10.1016/j.cortex.2017.06.002.

47. Frank MJ, Gagne C, Nyhus E, Masters S, Wiecki TV, Cavanagh JF, et al. fMRI and EEG Predictors of Dynamic Decision Parameters during Human Reinforcement Learning. J Neurosci 2015;35:485–94. 10.1523/jneurosci.2036-14.2015.

48. Loh DA, Hairi NN, Hairi FM, Peramalah D, Kandiben S, Hamid MAIA, et al. Effects of a Multicomponent Exercise and Therapeutic Lifestyle (CERgAS) Intervention on Gait Function in Lower-Income Urban-Dwelling Older Adults: A Cluster Randomized Controlled Trial. J Aging Phys Act 2023;31:531–40. 10.1123/japa.2022-0047.

49. Figueroa-Vargas A, Valenzuela R, Soto-Icaza P, Zamorano F, Larraín C, Silva C, et al. Deep White-Matter Pathways Mediate the Link Between Docosahexaenoic Acid (DHA) Status and Cognitive Performance in Adolescence. bioRxiv 2025:2025.11.20.689543. 10.1101/2025.11.20.689543.

50. Shahinfar H, Yazdian Z, Avini NA, Torabinasab K, Shab-Bidar S. A systematic review and dose response meta analysis of Omega 3 supplementation on cognitive function. Sci Rep 2025;15:30610. 10.1038/s41598-025-16129-8.

51. Kausel L, Michon M, Soto-Icaza P, Aboitiz F. A multimodal interface for speech perception: the role of the left superior temporal sulcus in social cognition and autism. Cereb Cortex 2024;34:84–93. 10.1093/cercor/bhae066.

52. Rosario MA, Kern KL, Mumtaz S, Storer TW, Schon K. Cardiorespiratory fitness is associated with cortical thickness of medial temporal brain areas associated with spatial cognition in young but not older adults. Eur J Neurosci 2024;59:82–100. 10.1111/ejn.16200.

53. d’Arbeloff T, Cooke M, Knodt AR, Sison M, Melzer TR, Ireland D, et al. Is cardiovascular fitness associated with structural brain integrity in midlife? Evidence from a population-representative birth cohort study. Aging (Albany NY) 2020;12:20888–914. 10.18632/aging.104112.

54. Olivo G, Nilsson J, Garzón B, Lebedev A, Wåhlin A, Tarassova O, et al. Higher VO2max is associated with thicker cortex and lower grey matter blood flow in older adults. Sci Rep 2021;11:16724. 10.1038/s41598-021-96138-5.

55. Erickson KI, Prakash RS, Voss MW, Chaddock L, Hu L, Morris KS, et al. Aerobic fitness is associated with hippocampal volume in elderly humans. Hippocampus 2009;19:1030–9. 10.1002/hipo.20547.

56. Frederiksen KS, Larsen CT, Hasselbalch SG, Christensen AN, Høgh P, Wermuth L, et al. A 16-Week Aerobic Exercise Intervention Does Not Affect Hippocampal Volume and Cortical Thickness in Mild to Moderate Alzheimer’s Disease. Front Aging Neurosci 2018;10:293. 10.3389/fnagi.2018.00293.

57. Festa F, Medori S, Macrì M. Move Your Body, Boost Your Brain: The Positive Impact of Physical Activity on Cognition across All Age Groups. Biomedicines 2023;11:1765. 10.3390/biomedicines11061765.

58. Chai Q-Y, Song A-Q, Zhao Q-Y, Shen Q-Q, Cui L. An ALE meta-analysis on the effects of neural changes due to exercise on executive function in a healthy population. Sci Rep 2025;15:34415. 10.1038/s41598-025-17431-1.

59. Tan F-M, Teo W-P, Leuk JS-P, Goodwill AM. Effect of habitual physical activity on motor performance and prefrontal cortex activity during implicit motor learning. PeerJ 2024;12:e18217. 10.7717/peerj.18217.

60. Stern Y. What is cognitive reserve? Theory and research application of the reserve concept. J Int Neuropsychol Soc 2002;8:448–60. 10.1017/s1355617702813248.

61. Wan L, Molina-Hidalgo C, Crisafio ME, Grove G, Leckie RL, Kamarck TW, et al. Fitness and exercise effects on brain age: A randomized clinical trial. J Sport Heal Sci 2025:101079. 10.1016/j.jshs.2025.101079.

62. Huang X, Zhao X, Cai Y, Wan Q. The cerebral changes induced by exercise interventions in people with mild cognitive impairment and Alzheimer’s disease: A systematic review. Arch Gerontol Geriatr 2022;98:104547. 10.1016/j.archger.2021.104547.

63. Esteban-Cornejo I, Stillman CM, Rodriguez-Ayllon M, Kramer AF, Hillman CH, Catena A, et al. Physical fitness, hippocampal functional connectivity and academic performance in children with overweight/obesity: The ActiveBrains project. Brain, Behav, Immun 2021;91:284–95. 10.1016/j.bbi.2020.10.006.

64. Cristi-Montero C, Hernandez-Jaña S, Zavala-Crichton JP, Tremblay MS, Ortega FB, Feter N, et al. Mentally active but not inactive sedentary behaviors are positively related to adolescents’ cognitive-academic achievements, a cross-sectional study — The Cogni-Action Project. Ment Heal Phys Act 2023;25:100561. 10.1016/j.mhpa.2023.100561.

65. Cristi-Montero C, Johansen-Berg H, Salvan P. Multimodal neuroimaging correlates of physical-cognitive covariation in Chilean adolescents. The Cogni-Action Project. Dev Cogn Neurosci 2024;66:101345. 10.1016/j.dcn.2024.101345.

66. Zhang Z, Chen Y, Yu Q, Li J, Zou L, Mavilidi MF, et al. A neurobiological taxonomy of sedentary behavior for brain health. Trends Neurosci 2025;48:853–64. 10.1016/j.tins.2025.09.002.

