## Supplementary_Material.docx for "Physical Activity Shapes Brain Structure, Function, and the Computational Mechanisms of Cognitive Control"

**Tables**

Table 1. Hippocampal subfields with greater volume in the active group

| **Hippocampus Structure** | **F-Statistic** | **p-value** | |
| --- | --- | --- | --- |
| *molecular_layer_HP-body_Lh:* | 7.26 | 0.0089 | ** |
| *CA3-body_Lh:* | 7.11 | 0.0096 | ** |
| *GC-ML-DG-head_Lh:* | 5.28 | 0.0247 | * |
| *CA4-head_Lh:* | 8.24 | 0.0055 | ** |
| *CA3-head_Lh:* | 8.35 | 0.0052 | ** |
| *GC-ML-DG-body_Lh:* | 5 | 0.0287 | * |
| *CA4-body_Lh:* | 5.63 | 0.0205 | * |

Table 2. Hippocampal shape differences

| **Structure** | **p_value** | | **r Rank BiSerial** | **Cohen_d** | **Effect Size** |
| --- | --- | --- | --- | --- | --- |
| *CA1-body_Lh* | 0.046 | * | 0.282 | 0.589 | Medium effect |
| *molecular_layer_HP-head_Lh* | 0.044 | * | 0.286 | 0.596 | Medium effect |
| *molecular_layer_HP-body_Lh* | 0.007 | ** | 0.384 | 0.831 | Large effect |
| *GC-ML-DG-head_Lh* | 0.016 | * | 0.341 | 0.724 | Medium effect |
| *CA3-body_Lh* | 0.01 | * | 0.364 | 0.781 | Medium effect |
| *GC-ML-DG-body_Lh* | 0.018 | * | 0.334 | 0.708 | Medium effect |
| *CA4-head_Lh* | 0.004 | ** | 0.41 | 0.9 | Large effect |
| *CA4-body_Lh* | 0.018 | * | 0.336 | 0.712 | Medium effect |
| *CA3-head_Lh* | 0.003 | ** | 0.427 | 0.944 | Large effect |
| *Whole_hippocampal_body_Lh* | 0.023 | * | 0.322 | 0.681 | Medium effect |
| *Whole_hippocampal_head_Lh* | 0.04 | * | 0.291 | 0.608 | Medium effect |
| *Whole_hippocampus_Lh* | 0.038 | * | 0.294 | 0.615 | Medium effect |

Table 3. Peak MSIT-related clusters

| **Cluster Index** | **Voxels** | **P** | **-log10(P)** | **Z-MAX** | **Z-MAX X (mm)** | **Z-MAX Y (mm)** | **Z-MAX Z (mm)** |
| --- | --- | --- | --- | --- | --- | --- | --- |
| 12 | 3823 | 5.85E-20 | 19.2 | 5.11 | -18 | -58 | 4 |
| 11 | 1301 | 1.66E-09 | 8.78 | 4.96 | 66 | -40 | 24 |
| 10 | 1232 | 3.76E-09 | 8.42 | 5.2 | -62 | -38 | 32 |
| 9 | 930 | 1.79E-07 | 6.75 | 5.67 | 46 | 24 | -8 |
| 8 | 664 | 6.50E-06 | 5.19 | 3.94 | -16 | -68 | 46 |
| 7 | 644 | 8.76E-06 | 5.06 | 4.93 | 10 | -2 | 8 |
| 6 | 547 | 3.83E-05 | 4.42 | 5.28 | 40 | 20 | -6 |
| 5 | 409 | 0.00037 | 3.43 | 4.69 | -48 | -74 | 20 |
| 4 | 393 | 0.00049 | 3.31 | 5.09 | 8 | 22 | 36 |
| 3 | 271 | 0.00465 | 2.33 | 5.59 | 16 | 16 | 66 |
| 2 | 202 | 0.0193 | 1.71 | 4.18 | -24 | 12 | 66 |
| 1 | 188 | 0.0262 | 1.58 | 4.66 | 46 | -76 | 14 |

Table 4. Boundary parameter clusters

| **Cluster Index** | **Voxels** | **P** | **-log10(P)** | **Z-MAX** | **Z-MAX X (mm)** | **Z-MAX Y (mm)** | **Z-MAX Z (mm)** |
| --- | --- | --- | --- | --- | --- | --- | --- |
| 6 | 1458 | 2.42E-10 | 9.62 | 4.66 | -22 | -60 | -4 |
| 5 | 1181 | 6.27E-09 | 8.2 | 4.64 | -12 | -40 | 44 |
| 4 | 1008 | 5.96E-08 | 7.22 | 4.44 | 66 | -4 | -18 |
| 3 | 369 | 0.00071 | 3.15 | 4.63 | -56 | -2 | -14 |
| 2 | 240 | 0.00842 | 2.07 | 4.34 | 12 | -30 | 52 |
| 1 | 188 | 0.0256 | 1.59 | 4.88 | 8 | 66 | 14 |

Table 5. Significant activity clusters for Active Boundary > Sedentary Boundary contrast.

| **Cluster Index** | **Voxels** | **P** | **-log10(P)** | **Z-MAX** | **Z-MAX X (mm)** | **Z-MAX Y (mm)** | **Z-MAX Z (mm)** |
| --- | --- | --- | --- | --- | --- | --- | --- |
| 2 | 210 | 0.0157 | 1.8 | 4.45 | -48 | -82 | 6 |
| 1 | 194 | 0.0222 | 1.65 | 4.49 | -16 | -60 | -14 |
